# Limitations of models for guiding policy in the COVID-19 pandemic

**DOI:** 10.1101/2022.06.30.22277091

**Authors:** Paul M McKeigue, Simon N Wood

## Abstract

At the outset of the COVID-19 epidemic in the UK, infectious disease modellers advised the government that unless a lockdown was imposed, most of the population would be infected within a few months and critical care capacity would be overwhelmed. This paper investigates the quantitative arguments underlying these predictions, and draws lessons for future policy.

The modellers assumed that within age bands all individuals were equally susceptible and equally connected, leading to predictions that more than 80% of the population would be infected in the first wave of an unmitigated epidemic. Models that relax this unrealistic assumption to allow for selective removal of the most susceptible and connected individuals predict much smaller epidemic sizes. In most European countries no more than 10% of the population was infected in the first wave, irrespective of what restrictions were imposed. The modellers assumed that about 2% of those infected would require critical care, far higher than the proportion who entered critical care in the first wave, and failed to identify the key role of nosocomial transmission in overloading health systems. Model-based forecasts that only a lockdown could suppress the epidemic relied on a survey of contact rates in 2006, with no information on the types of contact most relevant to aerosol transmission or on heterogeneity of contact rates.

In future epidemics, modellers should communicate the uncertainties associated with their assumptions and data, especially when these models are used to recommend policies that have high societal costs and are hard to reverse. Recognition of the gap between models and reality also implies a need to rebalance in favour of greater reliance on rapid studies of real-world transmission, robust model criticism, and acceptance that when measurements contradict model predictions it is the model that needs to be changed.

## Introduction

At the outset of the COVID-19 epidemic in the UK in early 2020, advice from the Scientific Advisory Group on Emergencies (SAGE) to the UK government relied on models that predicted that unlesss coercive restrictions of social contact (“lockdown”) were imposed without delay, the epidemic would infect most of the population within a few months and critical care facilities would be quickly overwhelmed [1, 2]. Together with reports from northern Italy that hospital services had been overwhelmed with cases requiring critical care, this led to an abrupt change of policy and the imposition of a lockdown starting on 24 March 2020. Subsequent management of the pandemic in the UK and other countries was based on suppressing transmission through restricting social contacts, vaccinating the entire adult population and a test and trace programme, rather than on focused protection of the vulnerable. Reversal, or even questioning, of these policies was difficult, in part because of implementation of the recommendation from SAGE’s behavioural science subgroup to attempt to overcome people’s own rational assessment of risk with “hard hitting emotional messaging” [3]. The rapid end of the first wave of the epidemic was attributed to lockdown, despite evidence by early May that infection rates had been in substantial decline before lockdown. As similar epidemic modelling approaches may be used to advise policy makers about emerging epidemics in the future, it is important to examine their limitations.

The advice that only a lockdown could prevent critical care capacity from being overwhelmed in the UK, presented in reports from modellers at Imperial College (IC) [1] and the London School of Hygiene & Tropical Medicine (LSHTM) [2] was based on four propositions: (1) in an unmitigated epidemic with basic reproduction number *ℛ*_0_ = 2.5, more than 80% of the population will be infected within a few months; (2) about 2% of those infected will require critical care; (3) to limit morbidity and mortality will require suppressing the epidemic by reducing the reproduction number below 1, rather than mitigation by shielding the vulnerable; (4) only a lockdown can reduce the reproduction number below 1. In a subsequent report the IC modellers asserted that these propositions applied globally [4]. This article examines the quantitative arguments underlying these propositions and the lessons that can be learned.

## Methods

The approach used by the IC and LSHTM modellers was based on the classic Susceptible-Exposed-Infected-Removed (SEIR) compartmental model formulated by Kermack and McKendrick [5] in 1927, in which the **transmission function** relating the infection rate to the proportion *S* of individuals remaining susceptible is assumed to be linear in *S* [6, 7]. This is equivalent to assuming that all individuals are equally susceptible and equally connected [8]. Kermack and McKendrick were well aware of this, stating in the abstract of their 1927 paper:

*In the present communication discussion will be limited to the case in which all members of the community are initially equally susceptible to the disease*.

From their model, Kermack and McKendrick derived an equation (Appendix Equation 3) for the final size of an unmitigated epidemic given the basic reproduction number *ℛ*_0_. With *ℛ*_0_ = 2.5, the herd immunity threshold is 60%, and the epidemic ends when 89% of the population have been infected.

The assumption that susceptibility and connectivity do not vary between individuals is unrealistic; relaxing this assumption to allow for heterogeneity gives rise to a **non-linear** transmission function [8–11] relating the infection rate to the proportion *S* remaining susceptible. This function is mathematically constrained to be non-negative, increasing, convex (second derivative non-negative), and smooth unless a discrete factor like vaccination has a large effect. Under these constraints the function can be approximated by a power law in which the infection rate is proportional to *S*^*λ*^, where *λ* is the **immunity coefficient**, taking values greater than or equal to 1 [9, 10] (see Appendix Equation 1). The classic model with no heterogeneity and a linear transmission function corresponds to *λ* = 1. Derivations of key mathematical results for SEIR models with heterogeneous mixing are given in the Appendix. For given values of the basic reproduction number *ℛ*_0_ and *λ*, the final size of an unmitigated epidemic can be calculated from Appendix Equation 2.

## Results

### 1. Predicted size of an unmitigated epidemic

Figure 1 shows that for any given value of *ℛ*_0_, the predicted final size of an unmitigated epidemic is smaller with a model that allows for heterogeneity than with a model with no heterogeneity. Figure 2 compares the predicted trajectory of the epidemic under a model with no heterogeneity (*λ* = 1) with the trajectory under a model with heterogeneity quantified by *λ* = 4, assuming *ℛ*_0_ = 2.5. With *λ* = 4, the herd immunity threshold is 20%, and the epidemic ends when 35% have been infected. For comparison, the values for the size of an unmitigated epidemic predicted by the IC modellers (who assumed *ℛ*_0_ = 2.4) and the LSHTM modellers (who assumed *ℛ*_0_ = 2.7) were respectively 81% and 85%. These values are slightly smaller than the calculated value for the size of an epidemic under a model with no heterogeneity, because the IC and LSHTM models allowed for slight heterogeneity attributable to variation of contact rates between age bands, equivalent to using *λ* = 1.2 in Appendix Equation 2.

**Fig 1.**
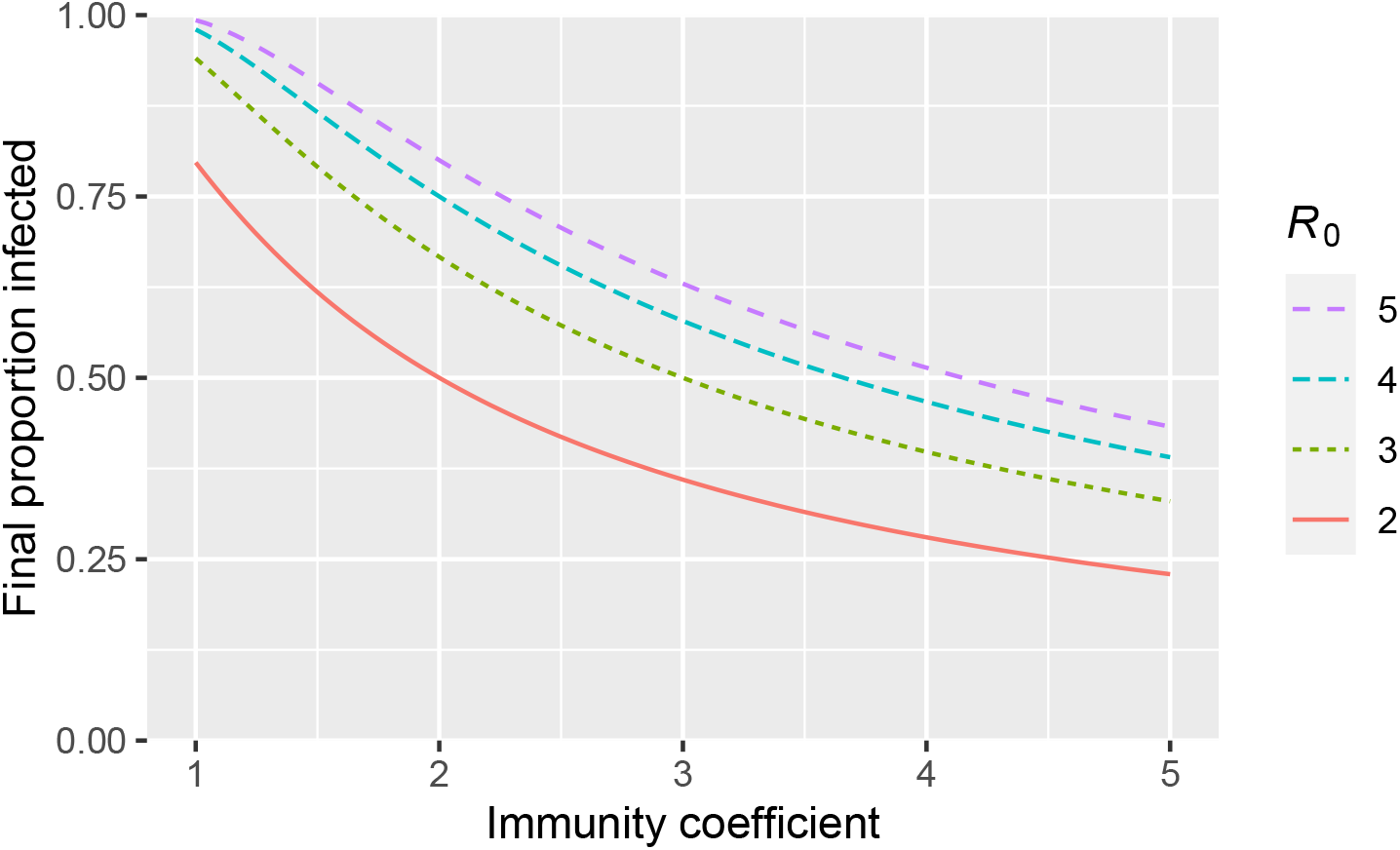
Relation of final size of unmitigated epidemic to immunity coefficient, for different values of the basic reproduction number *R*. See Appendix eqns. (2) and (3).

**Fig 2.**
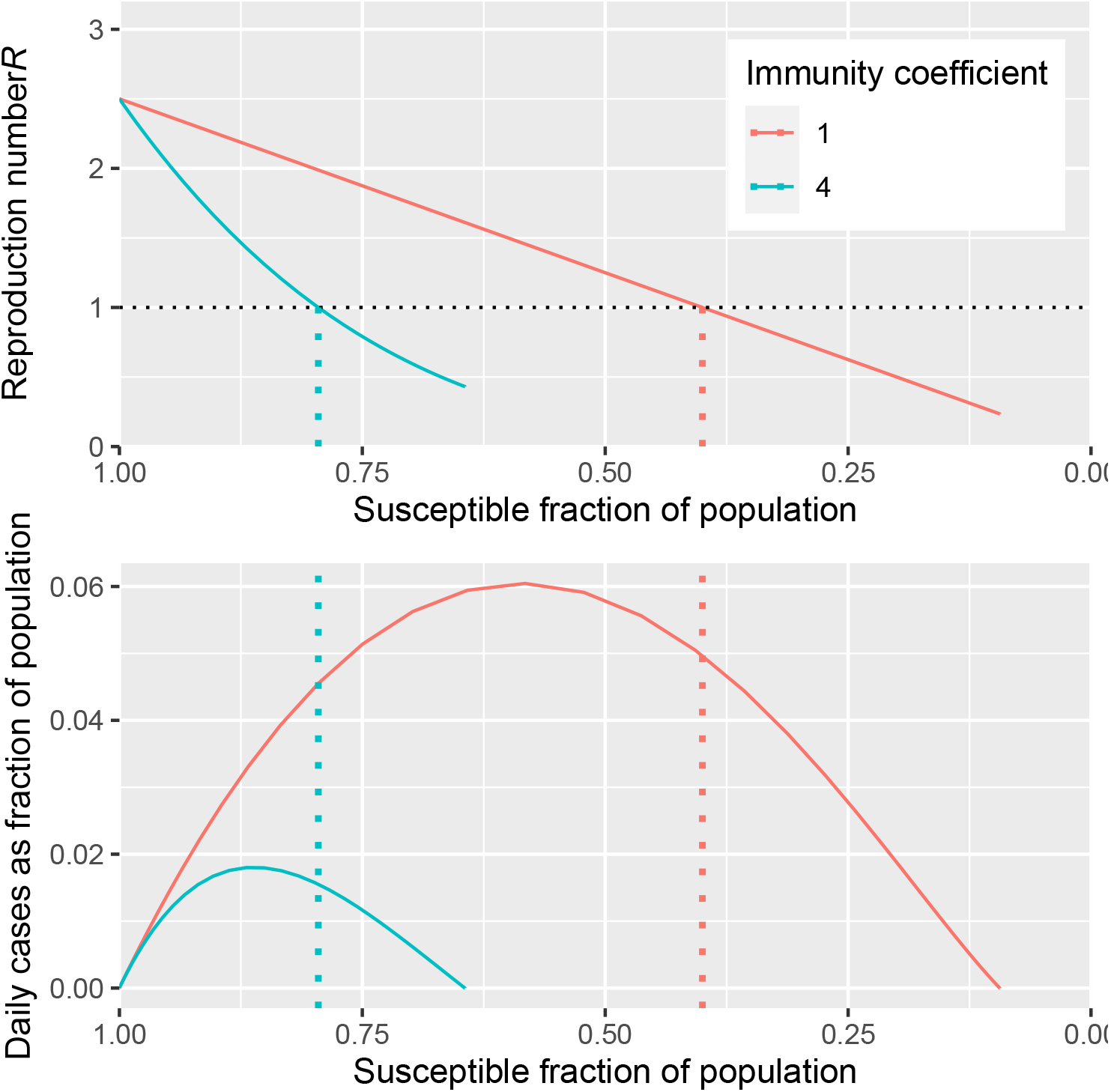
Relation of reproduction number and daily cases to the fraction of the population that is still susceptible, for immunity coefficients of one (no heterogeneity) and four. Vertical dotted lines are at one minus the herd immunity thresholds.

The underlying principle can be described without mathematics. Early in the epidemic, the most susceptible and the most highly connected individuals are selected for infection. Individuals who have persistently high contact rates with others – the hubs of the social network – are not only more likely to acquire infection but also more likely to transmit to others. As these highly susceptible or highly connected individuals are removed from the susceptible compartment, the infection will not spread so easily because the remaining individuals are more resistant to infection or have lower contact rates. The epidemic eventually kills itself by immunizing the most susceptible and the most highly connected individuals – the hubs of the social network. Varying infectiousness (for instance where a few “superspreaders” infect many people) does not affect the final size of the epidemic unless infectiousness is correlated with susceptibility.

#### Comparison of model-based predictions with the observed trajectory of the first wave

In most countries the proportion infected in the first wave up to August 2020 was less than 10% [12, 13]. Even in Iran, the first country outside China to be severely affected, seroprevalence was only 17% at the end of April 2020 [14]. As contact rates fell in all these countries, there is no example of an unmitigated first wave. Seroprevalence rates of more than 50% in the first wave have been reported only from the Amazonian region: 77% in blood donors in Manaus [15] in October 2020 and 70% in Iquitos in July 2020 [16]. The Manaus blood donors result appears to be an outlier, far higher than the adjusted seroprevalence of 29% in a cohort of Manuaus residents studied in October 2020 [15].

#### Quantifying heterogeneity

We can distinguish two sources of heterogeneity: biological susceptibility, and connectivity. There is indirect evidence for heterogeneity of biological susceptibility before the pandemic, based on studies showing that 20-50% of unexposed individuals had cross-reactive T cell responses to SARS-CoV-2 antigens [17]. Although later studies suggested that these cross-reactive T cell responses protect against infection [18, 19], no study has directly quantified the relation of infection risk to T cell reactivity in samples taken before the pandemic. In principle, the contribution of heterogeneity of connectivity to the immunity coefficient λ could be calculated from population-based surveys of the variance of contact rates between individuals. However as surveys of contact rates have recorded only a single day’s contacts in each individual [20, 21], no estimate of variance in persistent connectivity can be made since we can not separate between and within individual variability. The variance between age bands can be estimated however, and contributes an immunity coefficient of about 1.2 [9].

If biological susceptibility contributes to heterogeneity, we expect new epidemic waves to occur when new variants arise that overcome pre-existing resistance to the dominant strain. If connectivity contributes to heterogeneity, we expect new waves to occur when social networks are rewired, for instance at the beginning of a new school year. We can distinguish six epidemic waves in Britain up to early 2022: wave 1 starting in March 2020, wave 2 in September 2020, wave 3 in December 2020, wave 4 in May 2021, wave 5 in September 2021, and wave 6 in December 2021. Waves 2 and 5 were not obviously related to new variants, and coincided with the start of the school year. Waves 3, 4, and 6 were attributable to new variants: respectively Alpha, Delta and Omicron. This suggests that both sources of heterogeneity contributed to the wave-like pattern of the epidemic.

#### Learning the transmission function

Under variability in susceptibility or mixing rate models there is a one-to-one correspondence between the distribution of susceptibility or mixing and the transmission function. Hence we do not need to estimate the distribution of susceptibility or mixing rates if we can learn the transmission function from the trajectory of the epidemic. The reproduction number *ℛ* can be estimated from the rate of growth of the epidemic if the distribution of the serial interval – the time between successive cases in a chain of transmission – is known, without any other modelling assumptions [6]. In the UK the reproduction number is estimated to have fallen from about 3 at the start of the epidemic to about 0.5 in mid-April 2020, staying at around 0.8 until late summer [22, 23]. It is estimated that contact rates fell by 60% to 80% [12, 21, 24] after lockdown in comparison with the pre-pandemic period in the UK and other European countries.

Two studies have attempted to estimate immunity coefficients from the trajectory of the first wave, with models that allowed for changing contact rates [9, 11]. Using survey data to adjust for contact rates, the immunity coefficient λ was estimated as 2.9 for Scotland and England [9]. Using mobility data to adjust for contact rates, estimates of λ ranged from 4.1 to 4.7 in cities and states of the USA [11]. Although these results suggest that heterogeneity of connectivity is far more than is allowed for in the SAGE models, the results must be treated with considerable caution. It is statistically difficult to distinguish the effects of falling contact rates, reduced susceptibility among those remaining uninfected, seasonal effects on transmissibility, and the substantial modification of contact networks caused by lockdowns, because all these factors were changing at the same time. That said, [9] includes an extensive review of contact studies from which contact distribution parameters could be directly estimated: these suggest that the estimates were realistic.

Although published reports from SAGE examined the sensitivity of their forecasts to various modelling assumptions, sensitivity to the assumption of a linear transmission function was not included in these analyses. The rationale for ignoring unmeasured heterogeneity is not clear: in comments on social media, modellers have expressed scepticism that heterogeneity can be modelled without measuring susceptibility directly or relying on strong assumptions about the distribution of susceptibility in the population. However where modelling results are highly sensitive to a process, the difficulty of measuring that process is a poor excuse for ignoring it. In any case, it is not necessary to make strong assumptions about the distribution of susceptibility if we can learn the transmission function from the observed trajectory of the epidemic, although it is clear that attempting to do so may well increase the level of uncertainty that has to be acknowledged in the modelling results. Conversely, if no reliable estimate of the transmission function can be made from real-world observations, that would seriously undermine the case for using models to set policy at all.

### 2. Requirement for critical care

The MRC Biostatistics Unit estimates that about 10% of the English population was infected by the end of the first wave [25]. Over the same period about 0.1% of the population died from COVID-19 as underlying cause, giving an infection fatality rate of about 1%. This is close to the value of 0.9% assumed in the IC model at least for the first wave, though the infection fatality rate was heavily weighted by the high fatality rate in care home residents, who accounted for half of all fatal cases in Scotland [26]. However, the SAGE modellers’ assumptions about the ratio of cases requiring critical care to fatal cases were wide of the mark. The IC report assumed that the ratio of cases entering critical care (30% of 4.4% hospitalised) to fatal cases (0.9% of infections) would be 1.3 [1]. From the LSHTM report [2] the value assumed for the ratio of cases entering critical care to fatal cases at the peak of the epidemic can be calculated (from the projections of peak requirement of 200,000 critical care beds with average length of stay 10 days, and peak weekly deaths 57000) as 2.5. In Scotland during the first wave the actual ratio of cases entering critical care to fatal cases was 0.2 [26]. As critical care capacity was not exceeded, this low ratio of cases entering critical care to death was presumably because most of those who were destined to die from COVID-19 were assessed as unlikely to benefit from critical care because they were very frail and near the end of life.

Although the inability of health-care systems to cope with the first wave of the epidemic in Lombardy was interpreted as the result of failing to control population-wide transmission, a subsequent report noted that in Lombardy at this time “SARS-CoV-2 became largely a nosocomial infection” [27]. Nosocomial transmission accounts for a high proportion of severe cases because even if the clinically vulnerable can shield themselves from other sources of exposure, they cannot easily avoid exposure to hospital when they need medical care [28]. From 2 April 2020 onwards SAGE recorded increasing concern about nosocomial infection, but on 7 May noted that “Granular data are not yet currently available from PHE to fully understand transmission pathways in healthcare settings” [29].

### 3. Suppression or mitigation?

The optimal mitigation scenario considered by the IC modellers – 75% effective shielding of the 15% of the UK population who were aged over 70, combined with case isolation – was predicted to reduce mortality by two-thirds. The modellers argued that this would not be enough to keep the requirement for critical care within the limits of capacity, and that this would require reducing the reproduction number below 1 to suppress the epidemic.

Possibilities for a broader mitigation strategy based on risk-stratified protection of the vulnerable were not examined at this time. Based on the information available at the time about risk stratification, if all those past retirement age or with designated risk conditions had been advised to shield, at least 25% of the population would have been advised to shield initially, but this 25% at highest risk would have accounted for at least 85% of those at risk of fatal disease [30]. Those who are advised that they are at high risk are likely to restrict their contact rates voluntarily: focused protection of this group can be supported without coercion. For those in the high-risk group who were economically inactive, not living with economically active adults and able to live without personal care, only minimal support for shielding would have been required. Additional support for those who needed it – for instance because of exposure at work, carer responsibilities, or sharing a household with other adults who were likely to be exposed – could have been provided at far lower cost to society than population-wide restrictions on social and economic activity. When the NHS Volunteer Responders Programme was announced on 24 March 2020 with the aim of supporting 2.5 million individuals identified as clinically vulnerable to COVID-19, 750,000 volunteers came forward within the first six days before recruitment was paused [31]. By end of September 2020, though 385,000 volunteers had been approved and had used an app to register themselves as on duty, only 110,000 vulnerable individuals had been provided with support through this initiative.

### 4. Did suppression require lockdown?

The modellers used survey data on contact rates to predict how non-pharmaceutical interventions would change the reproduction number. If transmissibility and susceptibility are constant, the reproduction number is proportional to the average contact rate, or to the largest eigenvalue of the matrix of age-stratified contact rates [6]. The only survey of contact rates in the UK available in early 2020 was the POLYMOD survey in which 1012 individuals in the UK had recorded their contacts for a single day in 2005-6 [20]. As POLYMOD included only seven participants aged over 75, data on mixing in the most vulnerable age groups was wholly inadequate. The type of contact most relevant to aerosol transmission of respiratory viruses like SARS-CoV-2 – sharing a poorly ventilated indoor space [32] – was not recorded: contacts were recorded only as physical (skin-to-skin contact) or non-physical (two-way conversation only). On 14 April 2020 SAGE noted that “Evidence suggests that transmission risk outdoors is significantly lower than indoors” but this did not lead to any reassessment of forecasts based on survey data that did not distinguish indoor contacts from outdoor contacts [29]. As contacts were recorded only on a single day, the variance of persistent connectivity between individuals could not be estimated. It is doubtful whether any reliable predictions of the effect of interventions on the reproduction number could be made from such inadequate data.

The attribution of the fall in reproduction number below 1 between March and April 2020 in the UK to lockdown [33, 34] was based on very strong modelling assumptions: in a reanalysis with relaxation of these assumptions, the reproduction number was estimated to be falling rapidly before lockdown was imposed [22, 23]. This was supported by reconstruction of infection dates from symptom onset dates reported in the REACT-2 survey. A similar analysis showed that the reproduction number had been falling before the second lockdown in November 2020, and this was supported by direct estimates of infection rates from the ONS COVID-19 infection survey [23].

For Sweden, one of the few countries that did not impose a lockdown, the IC modellers predicted on 26 March that given *ℛ*_0_ = 2.7, mitigation with “social distancing of whole population” without a lockdown would lead to 34895 deaths from COVID-19 [4]. In the year up to the end of July 2020, the number of COVID-19 related deaths reported in Sweden was 5741 and the estimated number of excess deaths from all causes was 4329 [35]. These results, together with studies from other countries casting doubt upon the necessity of lockdown were available well before the second lockdown in the UK (early May 2020 for [22]) though final peer-reviewed publication was later [36–38].

## Discussion

The analyses above show that of the four propositions on which the recommendation for lockdown was based, one – the assumption that 2% of those infected would require critical care – was unequivocally wrong, and another – that mitigation through focused protection would not be effective in limiting morbidity and mortality – was not seriously questioned. The other two propositions – that in an unmitigated epidemic 80% would be infected, and that only a lockdown could suppress the epidemic – were reliant on strong but unrealistic modelling assumptions and weak data.

Unlike weather models, epidemic models were not developed as forecasting models, but rather as theoretical models to aid the understanding of epidemic processes. While weather models are continuously validated and re-trained against measurements (and related climate models have been subject to extensive refinement of component processes via ground-truth measurement and back-cast validation), such a firm grounding in empirical reality is elusive in epidemic modelling and impractical for models of a newly emergent disease. At the same time, unlike weather and climate models, epidemic models are not based on well-understood physics, readily susceptible to accurate mathematical description, but rather on complex human behaviour, which at best can be captured only crudely by tractable mathematical models. In this respect epidemic models perhaps better resemble economic forecasting models, but without the substantial streams of relevant data available to tune the latter.

It follows that epidemic models may well be suitable for answering such broad scale qualitative questions as ‘if we coercively suppress all social contact to the maximum extent possible will we drive *ℛ <* 1?’, but are simply not designed or calibrated for more nuanced quantitative questions, such as ‘how limited a set of restrictions would have a reasonable chance of avoiding severe health system overload?’.

Given these limitations, although it may be appropriate to use epidemic models for exploration of what would happen if reality resembled the model, this exercise needs to be accompanied by proper communication of uncertainties and a greater willingness to rapidly re-assess the models when data suggest poor calibration. For example, given the strong sensitivity of epidemic wave size to variance in mixing rates and susceptibility, no prediction or statement of uncertainty should be considered reliable if it fails to take this variability into account. It also follows that studies aimed at *measuring* such variability should be a priority if model predictions are to be used in a policy setting. For example, a cross-sectional or cohort study with regular testing for infection, like the ONS COVID-19 Infection Survey [39], could be used to quantify the heterogeneity of connectivity, if cohort members recorded diaries of contacts of different types. Similarly, if peripheral blood mononuclear cells were stored at baseline, such a cohort study could also be used to quantify the heterogeneity of susceptibility attributable to pre-existing T cell cross-reactivity.

The modelling reports at the outset of the epidemic in March 2020 failed to communicate that by relying on the unrealistic assumption of no unmeasured heterogeneity they were likely to overestimate the size of the epidemic, and that no reliable prediction of the effects of non-pharmaceutical interventions could be made without more information about the mode of transmission. This suggests that policy advice in future epidemics should rely less on *models*, with a greater priority given to the rapid establishment of high quality direct *measurement* studies, such as the ONS survey, and on shoe-leather epidemiology [40] investigating the key target variable: transmission to vulnerable individuals. SAGE minutes from April-May 2020 show that although they were alerted to evidence that transmission risk was higher indoors than outdoors, and that nosocomial transmission was increasing, this did not lead to any serious reconsideration of policies. A report from NERVTAG and EMG to SAGE on 22 July 2020 concluded that “it is possible that transmission through aerosols could happen where a person who generates significant amounts of virus is in a poorly ventilated space with others for a significant amount of time.” but gave only cursory attention to investigations of outbreaks that by this time had provided compelling evidence for aerosol transmission [41].

The choice between risk-stratified protection of the vulnerable and intervention to suppress population-wide transmission is not binary: clinically vulnerable individuals will need support to shield themselves whether or not attempts are made to suppress population-wide transmission. Both lockdowns and focused protection strategies divide the population into compartments with differing connectivity. With a lockdown, the high-connectivity compartment comprises essential workers and carers, and those who share a household with them. With focused protection of the vulnerable, the high-connectivity compartment would comprise those who are young, fit and not sharing a household with someone vulnerable. Sustainable measures to reduce transmission may also be preferable to “circuit-breaker” lockdowns [42] in that they avoid repeated disruption of social networks that may lead to network rewiring with new hubs. Although herd immunity induced by natural infection of highly susceptible or highly connected individuals is likely to be only transient, even this would gain time for development of vaccines or other measures to protect the more vulnerable.

Finally, the epidemic models treat all negative effects of interventions as externalities. This is not rational when the aim of epidemic management is to minimise all loss of life from the epidemic and associated disruption. It is also an example, along with ignoring heterogeneity, seasonality, nosocomial transmission and the aspects of transmissibility not captured by contact rates, of excluding effects that are difficult to quantify, irrespective of their importance for relevant outcomes. The consequent danger is of one-sided caution in the advice offered, where recommendations that are presented as precautionary with respect to the outcomes considered in the model, may be recklessly risky in terms of outcomes that are omitted. For example, although the Bank of England has estimated that the economic shock to the UK from the response to COVID is the largest in 300 years, and the effect of economic deprivation and economic shocks on mortality are well established [43], quantifying the effects of lockdown on years of life lost is not amenable to simple mathematical models of the kind used by the SAGE modellers. Given that the primary issue here is one of balance and fairness between controlling COVID and other health problems, the standard cost-effectiveness analyses usually employed to help ensure health equity offer an alternative approach. Although these still require some estimate of the life years saved by lockdowns (and other measures) simple bounding calculations are possible. These suggest that the cost per year of life gained by lockdowns was far higher than the threshold of £30,000 used by NICE to decide whether to recommend use of a new pharmaceutical intervention in the health service [44].

## Supporting information

Rmarkdown source code

## Data Availability

This paper uses only publicly available data. R code to generate the figures and numbers in the manuscript is provided in the Markdown source document.

## Declarations

### Declaration of conflicting interests

The authors declared no potential conflicts of interest with respect to the research, authorship, and/or publication of this article.

### Funding

The authors received no financial support for this work.

## Appendix

S(E)IR models with heterogeneity of susceptibility or mixing

The key results are concisely derived here. For more see [8–11].

### Epidemic size under heterogeneity in susceptibility versus linear transmission (no heterogeneity)

An S(E)IR model that allows susceptibility to vary between individuals can be written

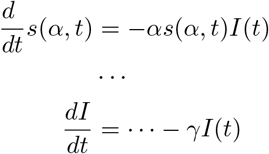

where … indicates details of model structure that do not change the results here. The positive transmission rate parameter *α* varies over the population (with susceptibility). Note that other authors write the transmission rate as the product of a population-level parameter *β* representing transmissibility, and an individual-level susceptibility parameter *ω* defined to have initial mean of 1 [8].

*s*(*α, t*)*dα* is the fraction of the population who are in the susceptible compartment at time *t* and have parameter value betwen *α* and *α* + *dα*. So if *S*_*t*_ is the fraction of the population who are susceptible at time *t* then *s*(*α, t*)*/S*_*t*_ is the probability density function of *α* within the susceptible compartment. Without loss of generality we define *S*_0_ to be 1. The first equation can immediately be integrated

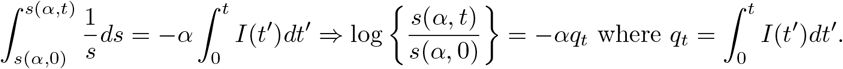

Hence *s*(*α, t*) = *s*(*α*, 0) exp(*αq*_*t*_). *q*_*t*_ has value 0 at *t* = 0, and increases with *t*.

Now suppose that the initial probability distribution for *α* is gamma(*k, ν*) with p.d.f. *s*(*α*, 0) = *ν*^*k*^*α*^*k*−1^*e*^−*αν*^*/*Γ(*k*). It follows directly that 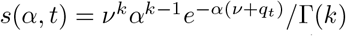. To obtain *S*_*t*_ we need to integrate *s*(*α, t*) over *α* (from 0 to ∞ where unspecified),

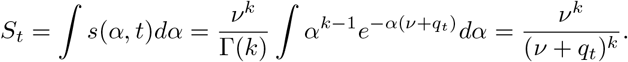

The second integral uses the identity

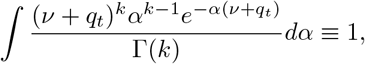

since the p.d.f. of a gamma(*k, ν* + *q*_*t*_) distribution integrates to one (like any p.d.f.). Similarly to find the time derivative of *S*_*t*_, we must integrate *α* out of the expression for the time derivative of *s*(*α, t*).

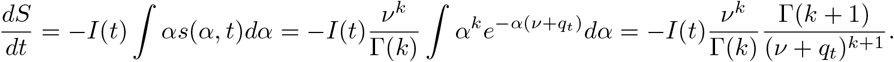

Again the second integral is obtained from the fact that a gamma(*k* + 1, *ν* + *q*_*t*_) p.d.f. must integrate to one. Using the identity *k* Γ(*k*) ≡ Γ(*k* + 1) and defining λ = 1 + 1*/k*, routine manipulation gives the transmission function as:

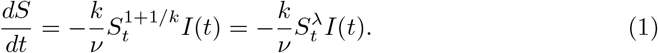

*ℛ*_0_, the expected number of new infections produced by an infectious individual at the start of the epidemic, is the mean *α* at time zero multiplied by the expected duration of infectiousness. The mean of a gamma(*k, ν*) random variable is *k/ν* so *ℛ*_0_ = *k/*(*νγ*). The final epidemic size is found by dividing (1) by the rate of removal *dR/dt* = *γI*, to get 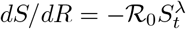. Rearrangement and integration gives

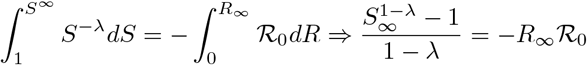

The final proportion *x* infected is *R*_∞_ = 1 − *S*_∞_, so routine re-arrangement then shows that *x* must satisfy

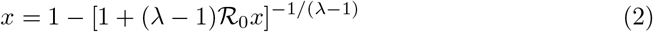

For λ = 1 the same approach shows that *x* must satisfy

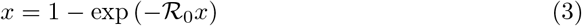

This is the limiting case in which *k* → ∞ and *ν* → ∞, while *k/ν* → a constant (for any meaningful model). For this to happen the variance of *α* must tend to zero - corresponding to no variability in susceptibility.

### Individual variation in mixing rates

The preceding model assumes that infectiousness is independent of susceptibility. This is a poor model for the case in which variability in transmissibility is related to variability in social contact rates. Therefore consider a model in which each individual has their own value of parameter *α*, and the transmission probability between individuals with parameter values *α* and *α*^*′*^ is proportional to *αα*^*′*^. Defining *I*(*α, t*) as the equivalent to *s*(*α, t*) for the infectious class, the susceptibility model is then modified to

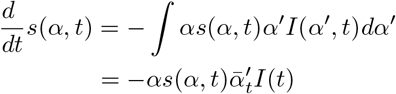

where 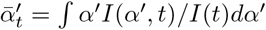. To proceed analytically, assume that the infectious state is short enough that, to a good approximation, the distribution of *α* in the infectious state at time *t* is given by the distributions of *α* in those first becoming infectious at time *t*. That is *I*(*α, t*) ∝ *αs*(*α, t*). Hence 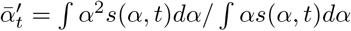 (the dividing integral is the normalizing constant for the p.d.f. of *α* in the *I* state).

Now redefining 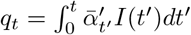, the algebra follows through exactly as in the variable susceptibility case so that 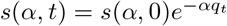, and again assuming a gamma(*k, ν*) initial *α* distribution we get

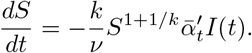

The need to evaluate 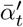 is new, but straightforward given that we again have an explicit gamma-like form for *s*(*α, t*). Using the same gamma integral tricks as above,

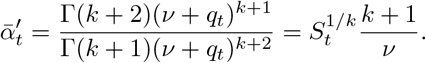

Clearly 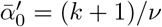, and the product of this, the initial expected value of *α*, and the expected residence time in the *I* compartment gives the basic reproductive number *ℛ*_0_ = *k*(*k* + 1)*/*(*ν*^2^*γ*). Hence

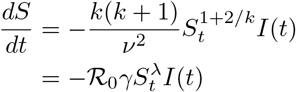

where now λ = 1 + 2*/k*. Clearly the equation to solve for the final epidemic size is again (2), all that has changed is the relationship between λ and *k*.

### Other distributions for susceptibility and contact rates

It is easy to work with any initial *α* distribution for which the moment generating function *ℳ* (*s*) = 𝔼 (*e*^*sα*^) exists (a very mild restriction). For a wide range of distributions the m.g.f., its inverse and derivatives are known and readily computed.

Under either model we have

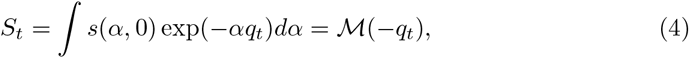

as the integral is simply the expectation defining the m.g.f. Differentiating the integral w.r.t. *q*_*t*_, and writing *ℳ* ′ (·) for the first derivative of *ℳ*, we also have,

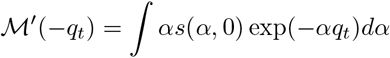

Hence, for the variable susceptibility model the transmission function is

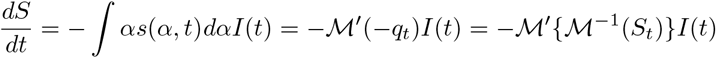

where *ℳ*^−1^ is the inverse function of *ℳ* and *q*_*t*_ = *ℳ* ^−1^(*S*_*t*_) follows directly from (4). So again we have a one-dimensional ODE. *h*(*S*) = *ℳ* ′ **{***ℳ* ^−1^(*S*)} is known as the *effective susceptible fraction*.

The variable mixing model is equally straightforward in the general distribution case. All that changes is that the right hand side of the ODE for *S*_*t*_ is again multiplied by the term 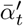. But differentiating the defining equation for *ℳ* (−*q*_*t*_) once more and substituting into the definition of 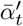 we obtain 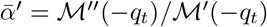, and hence the transmission function is

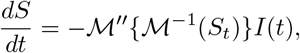

another one dimensional ODE with *h*(*S*) =*ℳ* ″ {*ℳ* ^−1^(*S*_*t*_)}.

[8] gives more detail, including examples for several distributions, and argues that the power transmission function obtained by assuming an initial gamma model is a good approximation more widely. However, once the original models are reduced from infinite dimensional to systems of a few readily computed ODEs in this way, they can readily be solved numerically to any desired accuracy, so approximation is somewhat superfluous.

## Notes

### Competing Interest Statement

The authors have declared no competing interest.

### Funding Statement

This study did not receive any funding

